# Estimating the Covid-19 Mortality Trend for Counties in the United States

**DOI:** 10.1101/2025.08.09.25333353

**Authors:** Kezia Irene, Sophia Qin, Sarita Pooranawattanakul, Mathew Samuel, Nebal Abu Hussein, Marie-Laure Charpignon, Adrien Carrel, Saketh Sundar, Leo Anthony Celi

**Affiliations:** Harvard Medical School, Boston, MA; Wellesley College, Boston, MA; Massachusetts Institute of Technology, Cambridge, MA

## Abstract

Covid-19 impacted counties in the United States differently in the first surge phase (Mar-Jun 2020). There are counties that have very high mortality (New York, NY) while there are counties that have low mortality rates (Grand, CO). We want to know if there is any predictor that could indicate why a county has higher or lower mortality rates. We also want to see if the social vulnerability index and other demographic factors play a role in the change of Covid-19 mortality. We have aggregated multiple datasets to find the Covid-19 predictors for counties in the United States. We also clustered these features using hierarchical clustering with dynamic time warping distance metrics and k-means clustering with exponential regression. According to K-means and Hierarchical Clustering using Dynamic Time Warping, Covid Mortality is divided into 5 different clusters. Using Elastic Net, we can conclude that crowding, income, stringency, access to the nursing homes, political leaning, percentage incarcerated, and being over 65 and impact to a higher covid Mortality. Having lower education being obese, being Hispanic, and being more distant from airports impact to a lower covid mortality. Social Determinants of Health: transportation, political leaning, income, and other ones that we tested for ARE significant in determining excess mortality.

## Introduction

There have been several studies assessing excess mortality during the COVID-19 pandemic across the United States and worldwide from 2020 to 2021^12345^, but there has not been much analysis done on contrasting the counties with the highest and lowest mortality rates in the United States. Moreover, there has been little investigation into why populations were more prone to excess mortality changes, whether that be positive or negative.

Excess mortality refers to the number of deaths caused by all crises that are beyond the number of deaths that we would expect to see in normal conditions ^6^. Excess mortality is measured by subtracting the number of reported deaths in a given time point in a crisis and the expected death when there is no crisis and can be seen in the equation below.

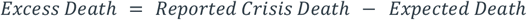

Research studies on the excess mortality associated with COVID-19 by the demographic group has shown that age, sex, and race showed differences in the mortality rates. Results were gathered from death certificates and used the historical all-cause death data to estimate the expected number of deaths, showing higher excess deaths in older populations and among Black adults^7^. Moreover, a national analysis of the COVID-19 epidemic and the study by the CDC both found that average deaths among Black populations were higher than white populations^8,9^.

One cross-sectional analysis study performed at a county level showed that the higher the social vulnerability score, the greater the risk of COVID-19 detection and death. Counties were identified in quartiles using the U.S. Center for Disease Control’s Social Vulnerability Index (SVI). The outcomes were positive tests and COVID-19 tests per capita. Outcomes were analyzed based on the counties’ characteristics and their heterogeneity in policies^10^.

Another systemic analysis research on COVID-19 related mortality demonstrated that the reported COVID-19 deaths between 2020-2021 were 5.94 million worldwide, but the estimated figure was 18.2 million as measured by excess mortality. To estimate excess mortality from Jan 1, 2020, to Dec 31, 2021, 6 prediction models were used to determine expected mortality. Excess mortality was calculated by subtracting deaths from COVID-19 with the expected mortality rates^11^.

The excess mortality for 2020 and 2021 is measured by fitting a regression model to each historical mortality data from 2015 to 2019. We analyzed the excess mortality for counties in the United States from March 2020 to June 2020 with features as shown in table 1.

**Table 1.** Features that possibly affect Covid-19 Mortality.

|  |  |
| --- | --- |
| • Date<br>• Confirmed deaths<br>• Recovered<br>• Tests<br>• Vaccines<br>• gathering restrictions<br>• school_closing<br>• workplace_closing<br>• cancel_events<br>• gatherings_restrictions<br>• transport_closing | • international_movement_restrictions<br>• information_campaigns<br>• testing_policy<br>• contact_tracing<br>• facial_coverings<br>• vaccination_policy<br>• elderly_people_protection<br>• government_response_index<br>• stringency_index<br>• containment_health_index<br>• Economic_support_index<br>• recreation_percent_change_from_baseline<br>• grocery_percent_change_from_baseline |

**Table 1.** Submission type, abstract length, and page length maximum for AMIA submissions.

| Data | Curating Organization | Source |
| --- | --- | --- |
| Counties | CDC Chronic Data | [12] |
| Nursing Homes | Centers for medicare and medicaid services | [13] |
| Incarceration Rates | Vera Institute | [14] |
| Race Demographic Survey | US census bureau | [15] |
| Policial Leaning | MIT | [16] |
| Distance to Airport | Department of Transportation | [17] |
| Stringency index | Oxford COVID-19 Government response tracker | [18] |

We want to see the trend and the differences in excess mortality because between 2020 and 2021. We want to see the predictor of an improvement in excess mortality, which counties have an improvement, and why different counties have different excess mortality.

Each county will be a data point and we plan to compare the trend for more than 100 counties. We will have the aggregate demographic information for each county, such as race, gender, ethnicity, and political leaning. Certain counties have more granular data than other counties, therefore we decided to use features owned by every county.

Our hypothesis is that overall there are factors that significantly impact positively and negatively the Covid-19 mortality during the first surge of Covid-19. We will analyze the correlation of excess mortality in counties in the US with their Social Vulnerability Index. We believe that more vulnerable counties will have higher mortality rates

In the next step, we plan to analyze the correlation of excess mortality in counties in the US with their Social Vulnerability Index (SVI). SVI refers to the communities’ ability to survive and thrive (resilience) when confronted with a crisis, such as a pandemic or natural disaster. We hypothesize that SVI will have a significant correlation with excess mortality in US counties. However, one of our main goals is to find the most significant SVI factors.

## Methods

We want to combine data from multiple sources to extend our features for each county. Then, we want to see which features matter to the mortality trend for each county using trend analysis. After that, we will do county trajectory clustering using K-means clustering and hierarchical clustering using dynamic time warping distance metrics, find the predictors that impact counties’ mortality rate using elastic net regression, and then finally find the importance of each predictor using the t-test. The method diagram can be seen in figure 1.

**Figure 1.**
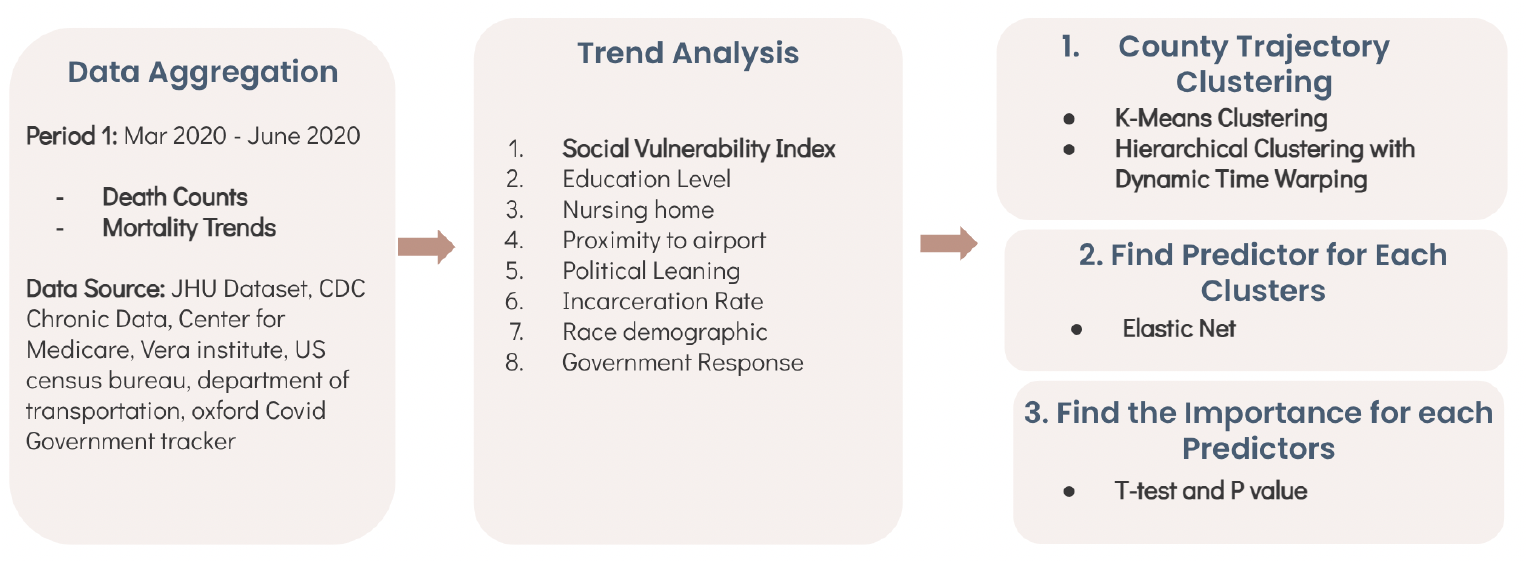
Methods overview

We combined data from multiple sources. For Covid mortality data, we curated the data from John Hopkins University dataset, county area data were curated from CDC chronic data, nursing homes were curated from centers for medicare and Medicaid services, incarceration rates were curated from Vera Institute, race demographic surveys were curated from US census bureau, political leaning was curated from MIT data, distance to the airport was curated from the department of transportation, and government response (stringency index) were curated from Oxford COVID-19 government response tracker. Full information about the data curation can be seen in table 1.

We used dynamic time warping algorithm to measure the similarity between 2 counties mortality time series per day. Dynamic time warping will calculate the optimal distance between two time series by calculating the distance between the next point in the comparison time series and the previous point in the comparison time series. After calculating the distance, a matrix will be created. Using this matrix, we then traverse through the lowest distance, therefore we could get the optimal match between two time series.

Hierarchical clustering is used to cluster the counties where we already had the distance matrix from doing the dynamic time warping. We use hierarchical clustering to make the hierarchy based on the dynamic time warping distance. Hierarchical clustering use euclidean distance to compare the position of one county to another.

We also compared our result with the K-means clustering result for the counties. In doing k-means clustering, we first determine the number of centroids by making an elbow plot. After that, we will find the lowest average distance between centroids and each point in the cluster.

In order to understand which SVI factors were significant in determining the mortality difference between the top 100 counties versus the bottom 100 counties, we performed statistical tests on Google Colab. Specifically, we calculated the t-statistic and p-values between the top 100 and bottom 100 for each factor. T-statistic is the ratio of the departure of the estimated value of a parameter from its hypothesized value to its standard error. The higher the t-value or more negative the t-value, the greater the confidence we have in the factor as a predictor. P-value is the level of marginal significance within a statistical hypothesis test, representing the probability of the occurrence of a given event. The closer the p-value is to zero the better.

## Results

We analyzed the mortality trends for COVID-19 between March 2020 and June 2020, where we can say it is the first surge of COVID-19 in the United States. By June 2020, we can see the mortality count in Figure 2. The darker the color for that county, the higher the number of mortality in that county by 30 June 2020.

**Figure 2.**
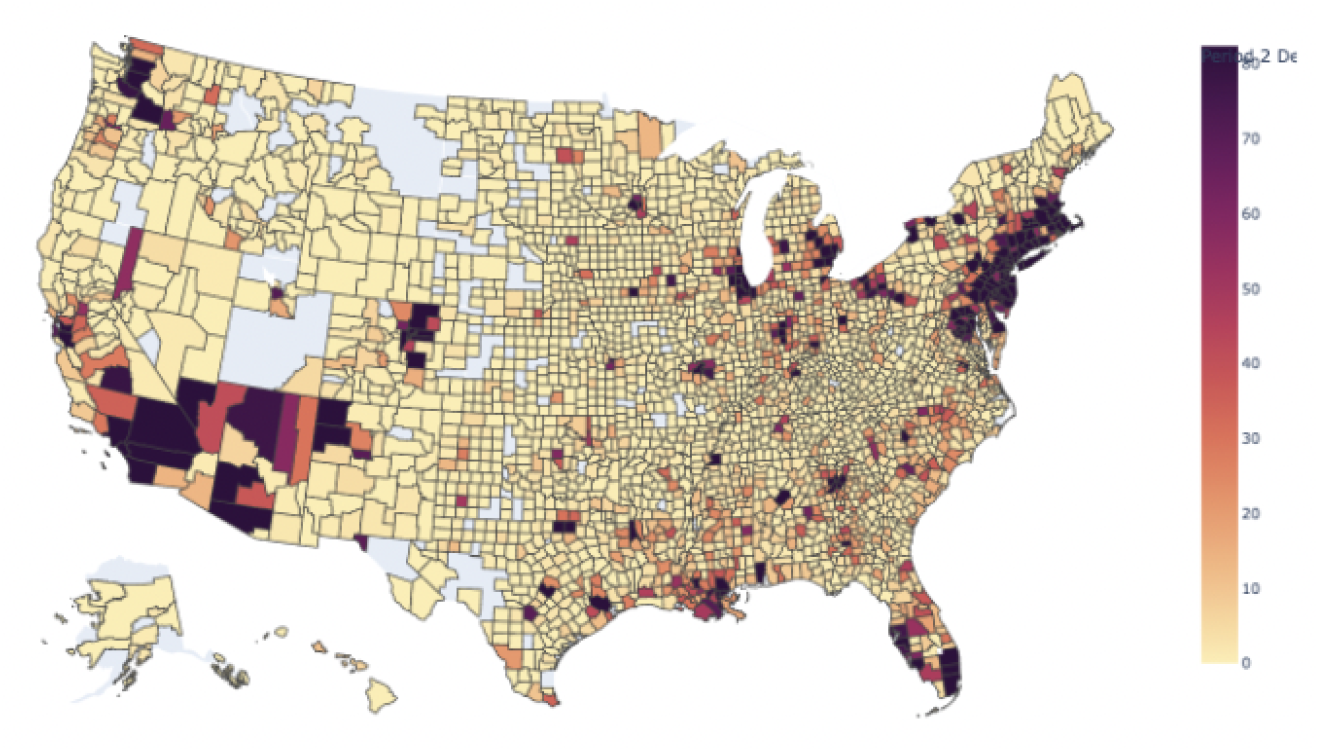
Mortality in June 2020

This data is good to give an overview of the situation in each county, but we are unable to find the predictor and the cluster for each county. Therefore we did a time series analysis. Figure 3 shows the overview of the time series analysis for 10 counties with the highest mortality and 10 counties with the lowest mortality.

**Figure 3.**
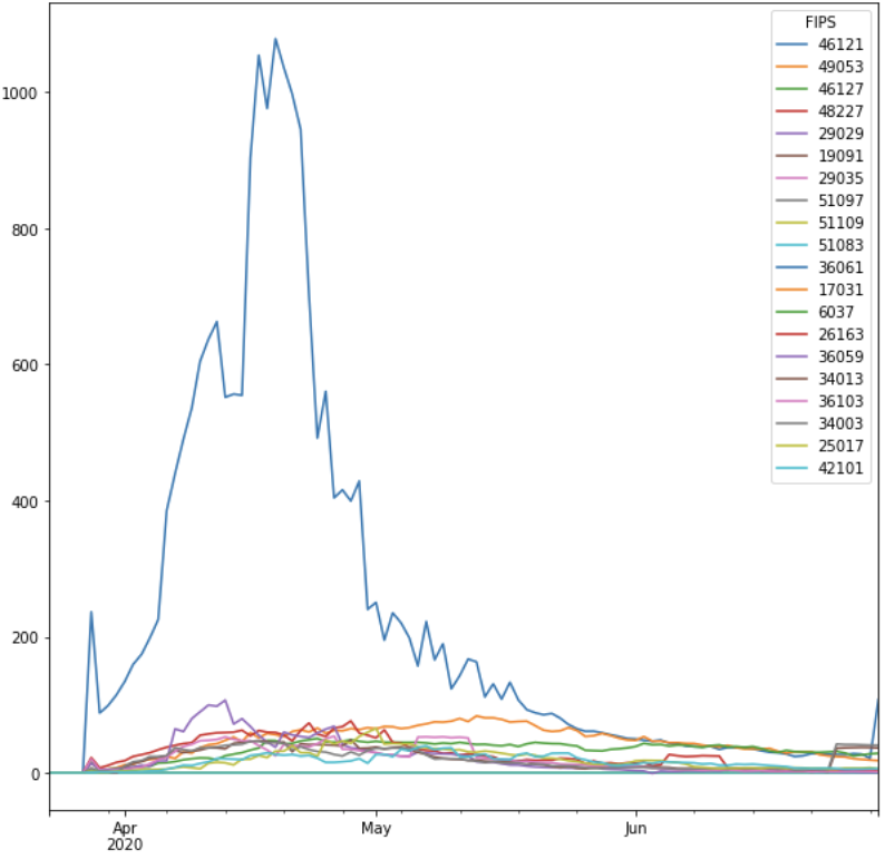
Time series of counties with highest and lowest mortality

After making the time series for each county, we did a hierarchical clustering using dynamic time warping metrics. We found that there are 5 optimal clusters using the hierarchical clustering algorithm, as seen in figure 4.

**Figure 4.**
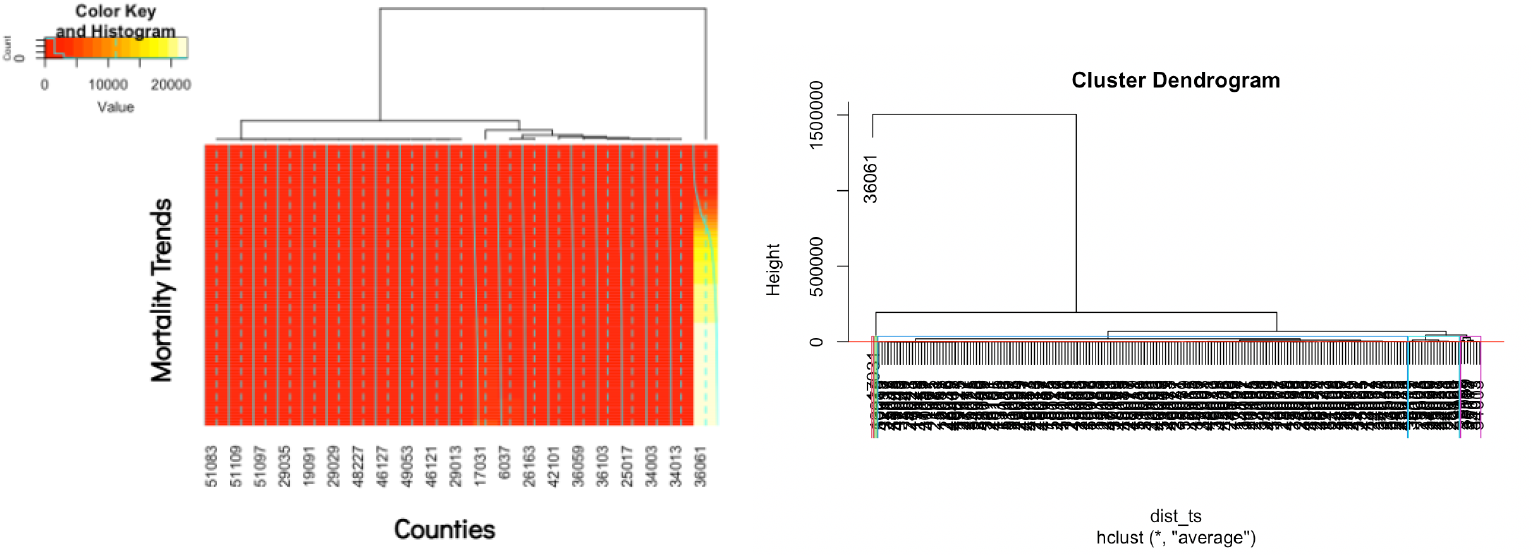
Hierarchical Clustering with Dynamic time warping distance metrics

**Figure 5.**
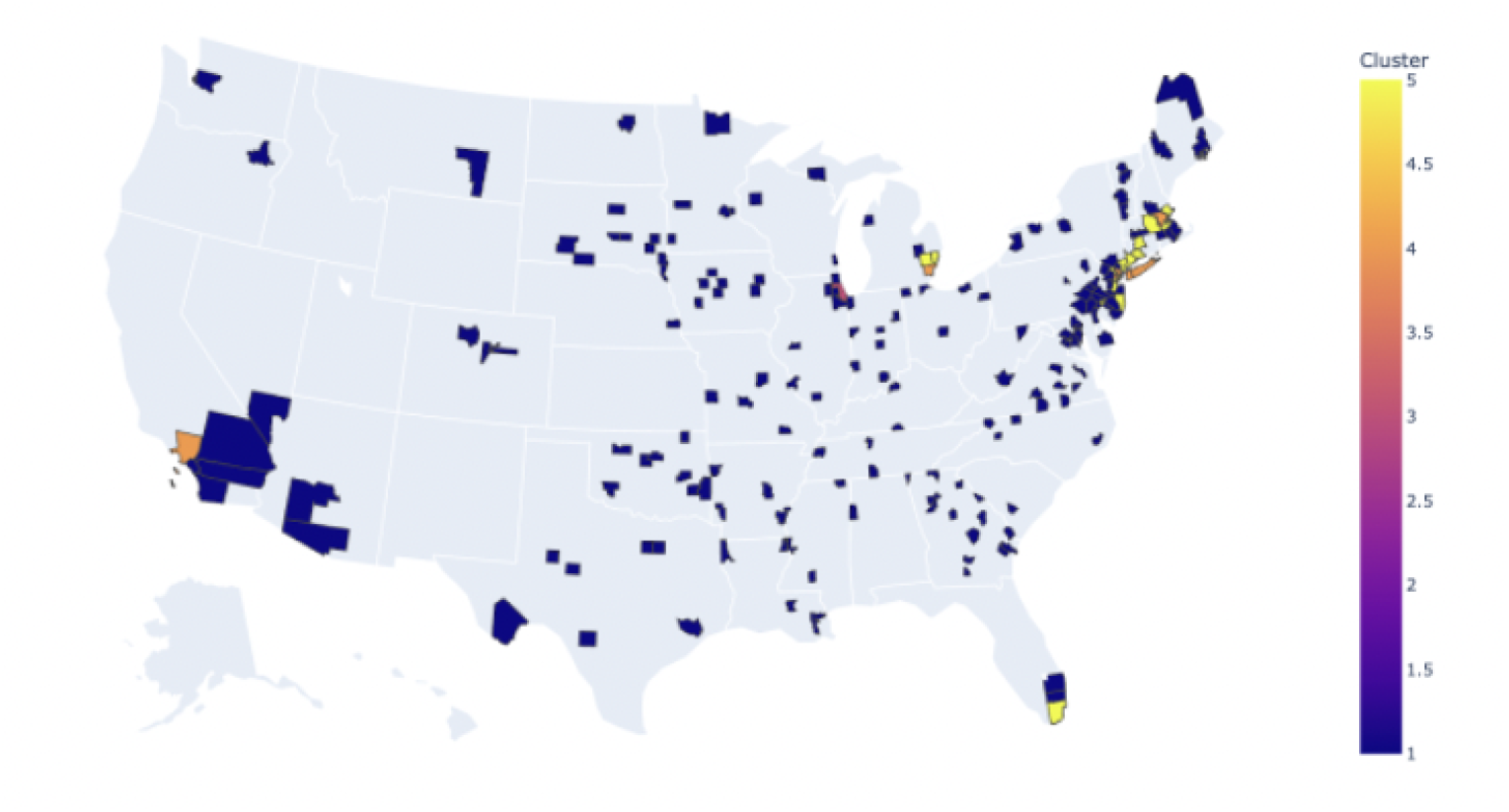
Hierarchical clustering map visualization

We then visualize the clustering of each county using map visualization. We can see the counties that are located near the coast are clustered together and have higher mortality trends, while counties that are located in the landlocked areas are clustered together and have lower mortality trends.

For the k-means clustering, we derived the time-series feature using the exponential regression function. Figure 6 shows the closeness and resemblance between the real trend and the exponential regression trends.

**Figure 6.**
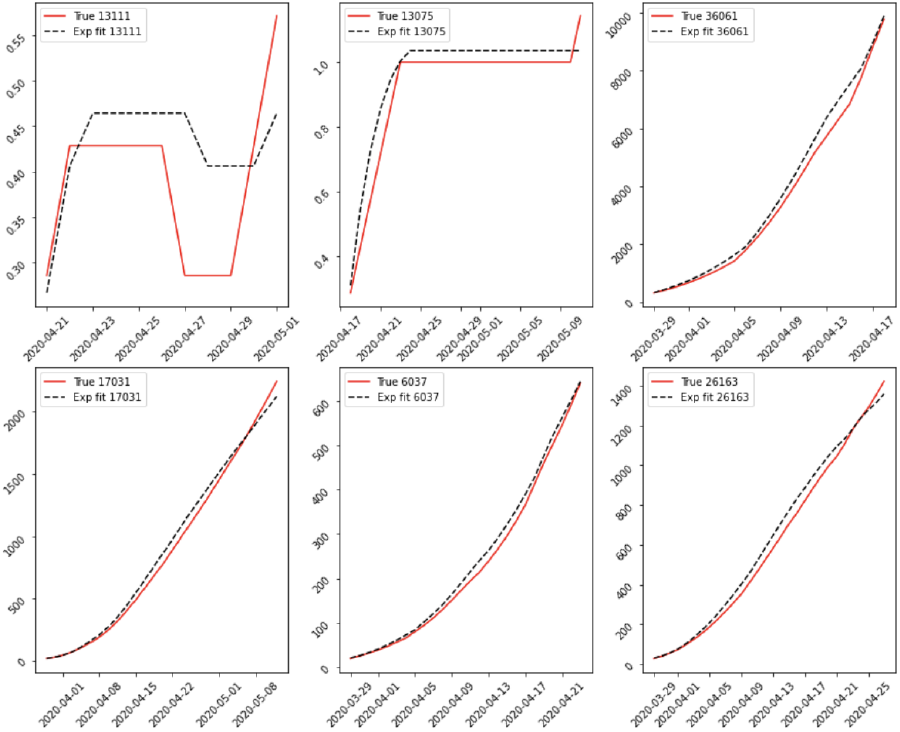
Exponential regression function compared to the actual trends

After calculating the elbow plot, we decided that for k-means clustering, the optimum number of clusters is 5 clusters. We visualize the clustering result using the county map that can be seen in Figure 7. We can see a similar pattern with the hierarchical clustering where counties that are located in the coast are grouped together while counties that are located in the midwestern are also grouped together.

**Figure 7.**
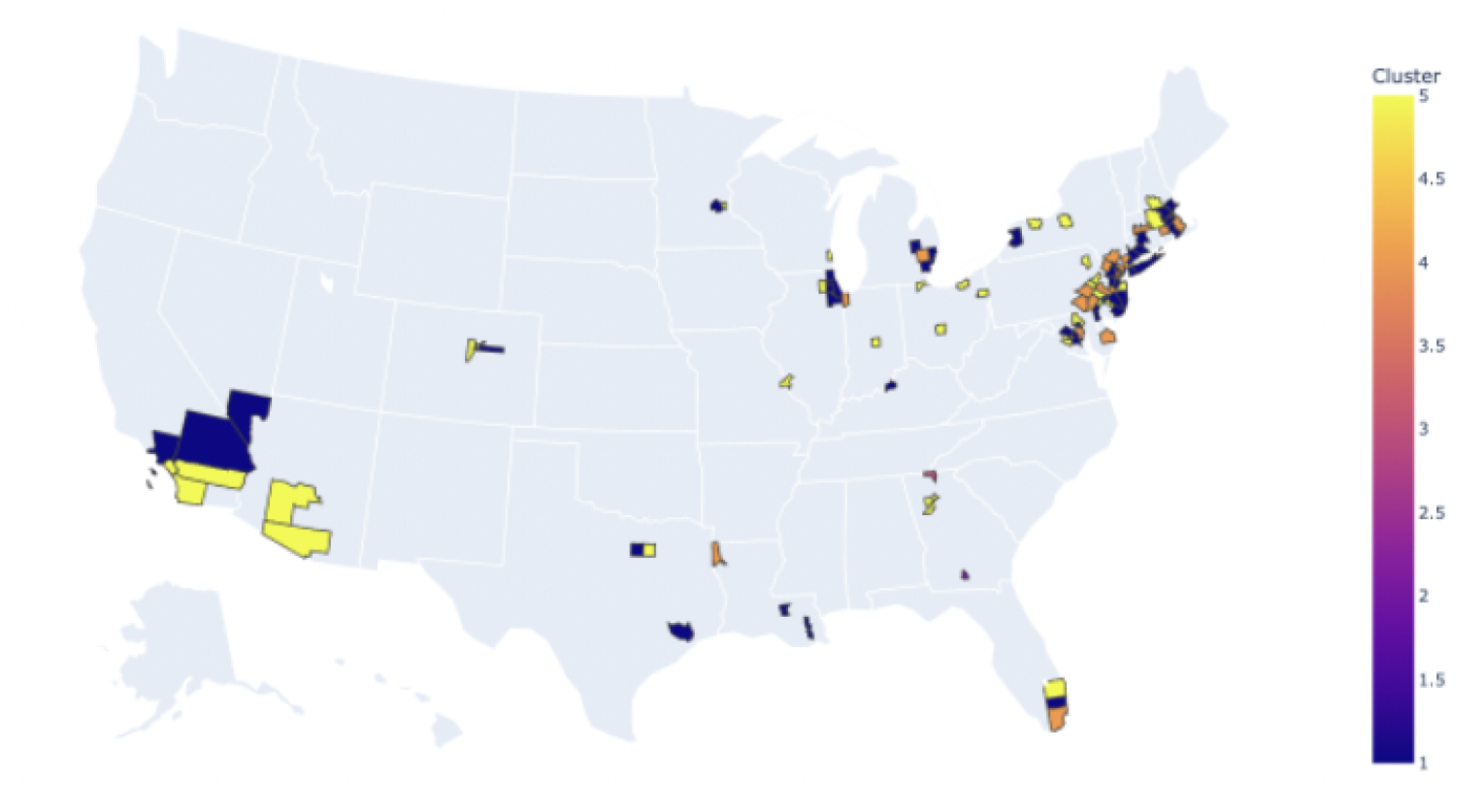
K-means clustering visualization.

**Figure 7a.**
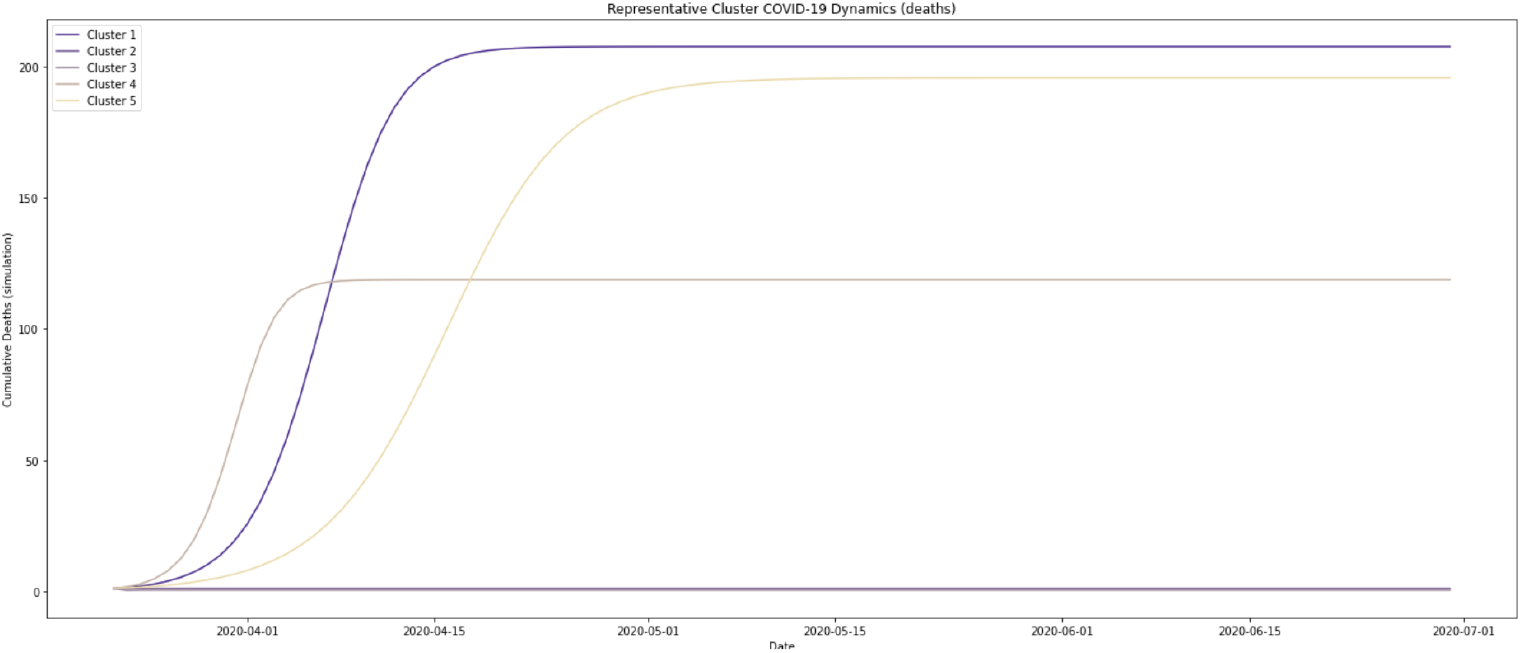
Trend Analysis per cluster

For the K-means clustering result, we also did a trend analysis to see the mortality trends within clusters. We can see that some clusters got higher rate of mortality in shorter time period while some other clusters got higher mortality rate but within more time.

We want to see the importance of each feature that we curated to predict COVID-19 trends in counties in the United States using the elastic net regression. We can see in figure 8 that more crowding, more income, right political leaning, and more incarcerated individuals lead to higher mortality rate. While the lower the obesity, the lower distance to the airport, and the lower people who age over 65 lead to a lower mortality rate.

**Figure 8.**
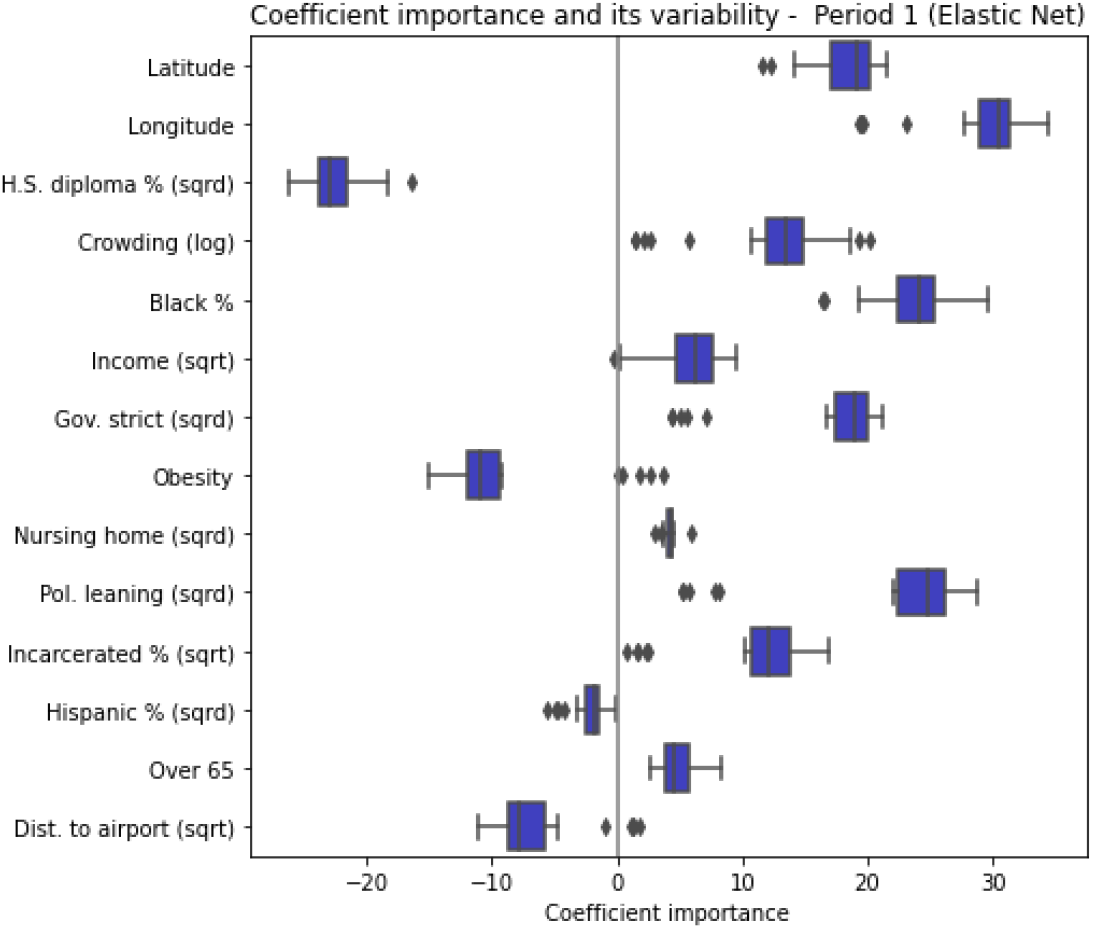
Feature Importance for Covid-19 mortality

We want to see if each feature has a significant different in average between the top 100 counties with highest mortality and the 100 counties with lowest mortality by June 2020 using T-test. The result can be seen in Table 2.

**Table 2.**

| Metric | P-value | T-statistic | Significant Factor |
| --- | --- | --- | --- |
| Proximity to airport | 0.000 | -13.155461 | yes |
| Political leaning | 0.000000 | -14.428287 | yes |
| Percentage white | 0.000000 | -6.300690 | Yes |
| Percentage black | 0.001575 | 3.206862 | Yes |
| Percentage asian | 0.000000 | 11.131106 | Yes |
| Percentage hispanic | 0.000000 | 7.255705 | Yes |
| Education | 0.000958 | 3.354986 | Yes |
| income | 0.000000 | 10.211132 | yes |
| Population density | 0.000292 | 3.690317 | Yes |

We created a merged data from various sources like different government collected censuses, then we ran some statistical tests to see which factors were statistically significant in differentiating between the top 100 and bottom 100 counties for excess mortality. We found that the significant factors were: proximity to airports, political leaning, percentage of white people, percentage of Asian people, percentage of black people, percentage of Hispanic people, education, income, and population density. Proximity to the airport meant the minimum distance to a top airport. By education, it meant the percentage of the population with a diploma greater or equal to a high school diploma. The income factor is the average annual income. Population density is equal to the total population in the county divided by the county area size. We were able to find the percentages of races in each county from the Race Demographic Survey collected by a government census. Political leaning was collected from the 2020 government election by calculating (votes_dem - votes_rep) /(votes_total). The more negative the number is the more Republicans, the less negative the more democratic the county is.

In conclusion, all the factors above were significant as their p-values were all less than 0.05. This means that in determining the difference between the top 100 and bottom 100 counties for mortality, these factors all played determining roles. The most significant values have a p-value of 0 and very large or very low t-stat values; this concludes the most significant factor is political leaning.

## Discussion

We tried to cluster 100 counties with the highest mortality count by June 2020 and 100 counties with the lowest mortality count by June 2020 to try to find the predictor of the mortality rate and see if there is any time-series-related pattern that can emerge from these total 200 counties. We found that using both k-means clustering and hierarchical clustering using dynamic time warping, we can divide the counties into four clusters. The counties that are located near the coast are clustered together and have a higher mortality rate. Counties that are located in landlocked areas are clustered together as well, where they have a lower mortality rate.

Some features are correlated with mortality rates as mentioned in the result part, and below is the reasoning on why some features are more correlated with mortality rate than other features:

### Correlation Between Political Leaning and Mortality Rates

The research topic, “The Changing Political Geography of COVID-19 Over the Last Two Years” by the Pew Research Center provided analysis and explanation of why political leaning could be one of the leading causes for higher or lower mortality rates. The main conclusions from this paper were that more Trump-leaning counties had lower rates of a public facility, restaurant dine-in, and public school closures. More conservative areas tended to have more lax COVID-19 policies. This explained the reason why after July 2020, pro-Trump areas had higher mortality. They deduced that there were more deaths in pro-Biden areas in the beginning most likely due to high population density in cities which tended to be more liberal. The population density was a determining factor for mortality rates in the beginning of the pandemic because there were no restrictions, mask mandates, vaccines, etc. yet.

### Correlation Between Race and Mortality Rates

A cross-sectional study, Racial and Ethnic Disparities in Population-Level Covid-19 Mortality (Journal of General Internal Medicine) using publicly reported Covid-19 mortality data. This study is focused on Black, Latinx, and White populations. They estimated the pooled effect of race on population-level mortality across states using inverse variance weighting. The report found strong relations between the Black race, Latinx ethnicity, and Covid-19 mortality rates. The report concluded that this made sense due to our country’s long-standing structural racism and the lack of COVID-19 information availability for populations of color.

### Correlation Between Proximity to Airport and Mortality Rates

In the report, “Geographic disparities in COVID-19 infections and deaths: The role of transportation,” from the Johns Hopkins Bloomberg School of Public Health, the relation between airports and COVID-19 is described. The main conclusions of this paper are that the number of COVID-19 deaths is positively related to proximity to and passenger volume of the nearest airport, the length of time between COVID-19 detection and death is also related to proximity to the airport, and finally, the numbers of deaths and cases were positively correlated with the number of airports. This paper urged for more stringent measures and tracing in the future to prevent higher risks of outbreaks for those who live and work near airports.

### Correlation Between Education and Mortality Rates

This study, “Variation in COVID-19 Mortality in the US by Race and Ethnicity and Educational Attainment,” by the Harvard T.H. Chan School of Public Health hints that educational attainment is tied to race and ethnicity. The report studied 219.1 million adults aged 25 years or older and found a correlation between racial groups, highest educational level attained, and COVID-19 mortality rates. For example, non-Hispanic White men with the least education died at a rate similar to the rates of college-educated non-Hispanic Black men. Ultimately, it concluded that higher educational attainment among racial and ethnic minority populations was insufficient to overcome racial and ethnic inequality–although education level affected mortality rates, race and ethnicity had a higher significance.

### Correlation Between Income and Mortality Rates

The study, “Association Between Income Inequality and County-Level COVID-19 Cases and Deaths in the US,” by Stanford University found that the pandemic highlighted the inequality across different socioeconomic classes. Their main goal was to evaluate the correlation between county-level income inequality, measured by the Gini coefficient, and county-level COVID-19 case and mortality rate across United States counties at different time periods across 2020 and 2021. The study found that counties with a lower average income had less access to personal protective equipment, COVID-19 testing, guidance on COVID-19 non-pharmaceutical interventions, education, and also, less vaccine acceptance.

### Correlation Between Population Density and Mortality Rates

In this ecological study, researchers from Universidade Federal de Sergipe investigated the relationship between population density and COVID-19 rates in a county-level analysis. This study of Brazil found it is a country with social inequalities: millions of people living in highly dense communities, with precarious housing conditions and poor sanitation will increase the risk of SARS-CoV-2 infection for those who live in highly dense areas. This study has been compared to those of Japan, India, Italy, the United States, and China, and similar conclusions have been reached. Studies on population density and COVID-19 mortality rates are important as they help determine the planning of policy and medical resources, and is a related measure to social distancing capacity.

### Social Vulnerability Index Discussion

In our samples of counties, the highest mortality rate was recorded in New York County: 22,521 deaths between 3/22-6/30/2020. The county with the lowest mortality recorded was in Grand Colorado. The relationship between social vulnerability and mortality rate was not stationary but varied between U.S. counties.These spatially variable findings contribute to other studies that demonstrated the nonuniform distribution of social vulnerability to U.S. disasters.

There is a clear relationship between the Social Vulnerability Index and mortality rates. It is found that 44 million adults are underinsured due to their socioeconomic class, race, income, etc. Racial disparity is clear as 2 million Americans lack running water in their homes, with Native Americans 19 times more likely and African Americans and Hispanics twice as likely to lack indoor plumbing than whites. Pandemics can profoundly and inequitably affect the health of socially vulnerable populations.Such events expose inequities in areas including access to quality housing and education as well as issues of economic and environmental justice that create conditions that make it difficult to maintain health.

We recognize that our study has limitations. Some limitations of our study include that our project only analyzed the top 100 counties with the highest mortality rate and the lowest 100 counties with the lowest mortality rate. Our project also only analyzed phase 1 of the Covid-19 surge in the United States (March - June 2020). In addition, in the future, we will test more factors from the Social Vulnerability Index like gender, employment, ownership of a vehicle, age, and immigrant status. In the future, we also hope to use our research more and apply it to predicting future pandemic outcomes.

## Conclusion

We have aggregated multiple datasets to find the Covid-19 predictors for counties in the United States. We also clustered these features using hierarchical clustering with dynamic time warping distance metrics and k-means clustering with exponential regression. According to K-means and Hierarchical Clustering using Dynamic Time Warping, Covid Mortality is divided into 5 different clusters. Using Elastic Net, we can conclude that crowding, income, stringency, access to the nursing homes, political leaning, percentage incarcerated, and being over 65 and impact to More Covid Mortality. Having lower education being obese, being Hispanic, and being more distant from airports impact to a lower covid mortality. Social Determinants of Health: transportation, political leaning, income, and other ones that we tested for ARE significant in determining excess mortality.

## Data Availability

All data produced in the present work are contained in the manuscript

## References

1. Lukowsky, L.R.; Der-Martirosian, C.; Dobalian, A. Disparities in Excess, All-Cause Mortality among Black, Hispanic, and White Veterans at the U.S. Department of Veterans Affairs during the COVID-19 Pandemic. Int. J. Environ. Res. Public Health 2022, 19, 2368. 10.3390/ijerph19042368

2. Troy Quast, PhD ; and Ross Andel, PhD, Excess Mortality Associated With COVID-19 by Demographic Group: Evidence From Florida and Ohio. sagepub.com/journals-permissions DOI: 10.1177/00333549211041550 journals.sagepub.com/home/phr

3. Emilio A. L. Gianicolo, Antonello Russo, Britta Büchler, Katherine Taylor, Andreas Stang, Maria Blettner. Gender specific excess mortality in Italy during the COVID‐19 pandemic accounting for age: European Journal of Epidemiology (2021) 36:213–218 10.1007/s10654-021-00717-9

4. New York City Department of Health and Mental Hygiene (DOHMH) COVID-19 Response Team; Preliminary Estimate of Excess Mortality During the COVID-19 Outbreak — New York City, March 11–May 2, 2020. Weekly / May 15, 2020 / 69(19);603–605

5. Janine Aron and John Muellbauer; Measuring excess mortality: the case of England during the Covid-19 Pandemic, INET@Oxford, updated and revised version, 18 May 2020

6. https://ourworldindata.org/excess-mortality-covid

7. Excess Mortality Associated With COVID-19 by Demographic Group: Evidence From Florida and Ohio. (https://journals.sagepub.com/doi/10.1177/00333549211041550

8. Rossen, LM., Branum, AM., Ahmad, FB., Sutton, P., Anderson, RN. Excess deaths associated with COVID-19, by age and race and ethnicity—United states, January 26–October 3, 2020. MMWR Morb Mortal Wkly Rep. 2020;69(42):1522–1527.doi:10.15595/mmwr.mm6942e2

9. Centers for Disease Control and Prevention. Excess deaths associated with COVID-19. January 17, 2021. Accessed February 12, 2021. https://www.cdc.gov/nchs/nvss/vsrr/covid19/excess_deaths.htm

10. County-Level Association of Social Vulnerability with COVID-19 Cases and Deaths in the USA (10.1007/s11606-020-05882-3)

11. Estimating excess mortality due to the COVID-19 pandemic: a systematic analysis of COVID-19-related mortality, 2020–21 (10.1016/S0140-6736(21)02796-3

12. https://chronicdata.cdc.gov/500-Cities-Places/PLACES-Local-Data-for-Better-Health-County-Data-20/swc5-untb

13. https://data.cms.gov/covid-19/covid-19-nursing-home-data

14. https://github.com/vera-institute/incarceration-trends

15. https://www.census.gov/newsroom/press-releases/2019/acs-5-year.html

16. https://github.com/TheUpshot/presidential-precinct-map-2020

17. https://data.transportation.gov/Aviation/International_Report_Passengers/xgub-n9bw

18. https://github.com/OxCGRT/covid-policy-tracker/blob/master/data/OxCGRT_latest.csv

